# Effects of acute or cumulative 8-hour sleep loss on simulated driving in a naturalistic, monotonous night-time setting: a randomized, controlled, crossover trial

**DOI:** 10.64898/2026.01.08.26343648

**Authors:** Stefan Lakämper, Michael Scholz, Thomas Kraemer, Hans-Peter Landolt, Kristina Keller

## Abstract

Sleep loss is a major contributor to traffic accidents, yet the differential effects of acute versus cumulative 8-hour sleep loss on vehicle control remain poorly quantified. This study leveraged tightly controlled sleep manipulation together with highly immersive driving simulation to directly compare sleep-loss modes under naturalistic driving conditions.

To evaluate subjective sleepiness, driving performance and vigilance, healthy male participants completed a randomized within-subject crossover sleep design, comprising normal sleep, cumulative sleep restriction (four nights with 2 h reduced sleep per night), and acute total sleep deprivation (one night without sleep), each followed by a simulated nighttime-to–early-morning driving protocol segmented into modules. Linear mixed-effects models evaluated condition effects and time-on-task dynamics.

Acute total sleep deprivation produced pronounced impairments in vehicle control. Most prominently, lateral control deteriorated rapidly under acute deprivation, with effect magnitudes comparable to those reported at high blood alcohol concentrations. Steering instability and integrated driving performance showed convergent deterioration. In contrast, cumulative sleep restriction resulted in smaller and less consistent changes, while vigilance task performance and speed variability were comparatively preserved across all conditions.

Despite increased subjective sleepiness, cumulative sleep loss provoked relatively small changes in driving performance, whereas acute sleep deprivation produced a disproportionate risk to driving safety by selectively and rapidly degrading operational vehicle control. These findings, obtained under ecologically relevant sleep and driving conditions, underscore functional and potentially underlying physiological differences between acute and cumulative sleep loss and, practically, highlight the importance of distinguishing between them when considering sleep-loss–related driving impairment and safety risk.

**Graphical Abstract:** 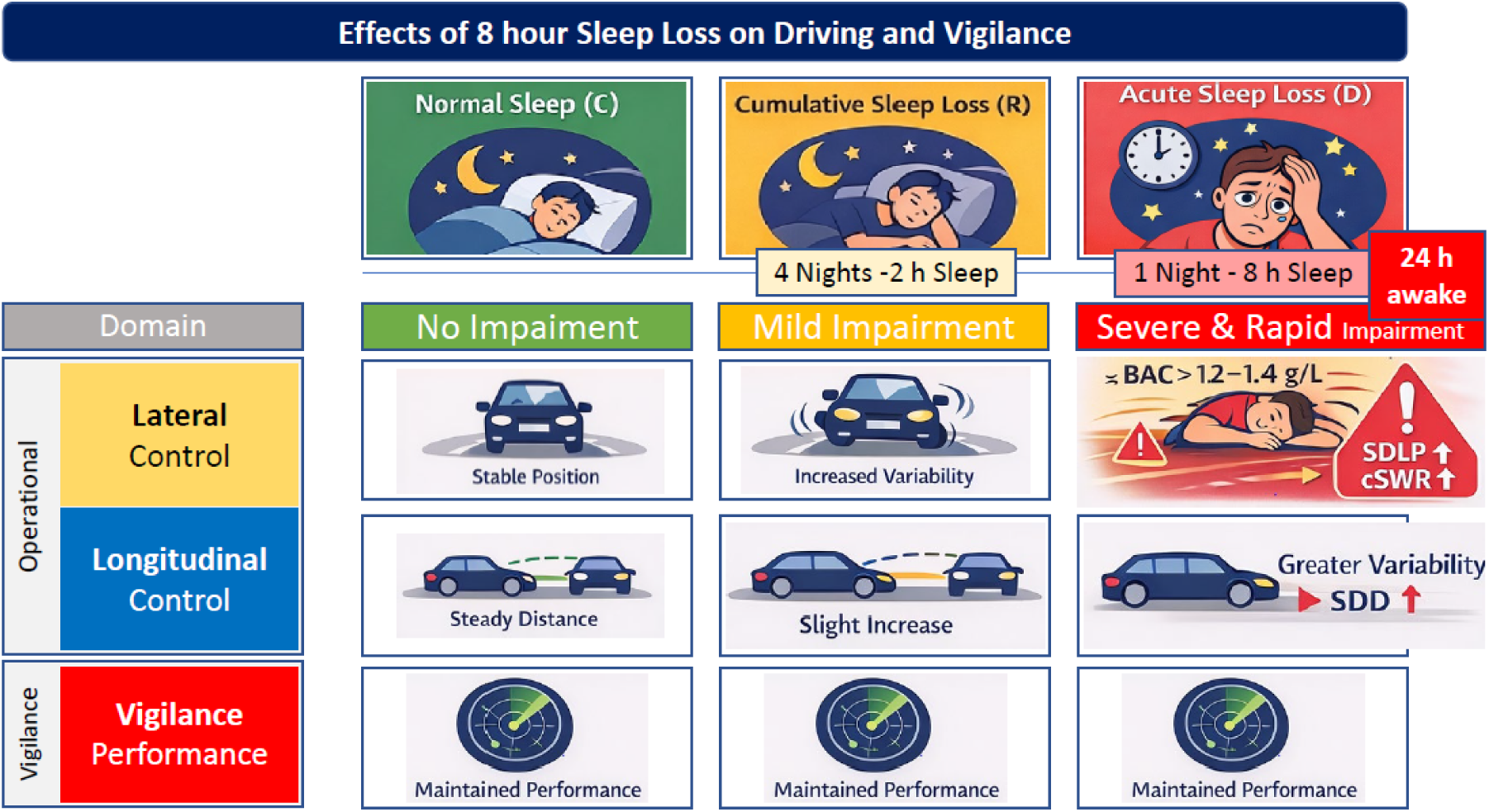

**Clinical Trials – name, URL and registration:** *Clinical Trial name:* Study of Identification of Metabolomics-based Sleepiness Markers for Risk Prevention and Traffic Safety - ClinicalTrials.gov Identifier NCT05585515, released on 18.10.2022, https://clinicaltrials.gov/study/NCT05585515; Swiss National Clinical Trial Portal SNCTP000005089, registered on 12.08.2022, https://www.humanforschung-schweiz.ch/de/studiensuche/studien-detail/58930

*IRB Statement:* This study was approved by the local ethics committee (Kantonale Ethikkommission Zürich, reference number 2022–01273). No identifying images or other personal or clinical details of participants are presented here or will be presented in reports of the trial results. Informed consent materials are available from the corresponding author on request. We confirm all methods were performed in accordance to the Declaration of Helsinki and its later amendments.

**Statement of significance:** Sleep loss is common, but not all forms of sleep loss affect driving in the same way. This study shows that a single night without sleep and several nights of restricted sleep have fundamentally different consequences for vehicle control, despite producing significant feelings of sleepiness. By combining controlled sleep manipulation with immersive, ecologically relevant driving assessment, the work reveals that acute sleep deprivation uniquely disrupts the ability to maintain stable control of a vehicle, whereas cumulative sleep loss produces more limited performance changes. These findings challenge assumptions based solely on subjective sleepiness and highlight the need to distinguish between sleep-loss patterns when evaluating driving safety. Future work should address how drivers recognize and respond to these distinct forms of impairment.

## Introduction

Insufficient or poor-quality sleep greatly increases the risk of motor vehicle accidents, regardless of whether the cause is a medical condition such as obstructive sleep apnea, restless leg syndrome, periodic limb movement disorder or narcolepsy, or results from social or occupational factors such as shift work, stress, or a voluntary sleep debt.

A substantial number of sleepiness-related accidents is likely unreported (*dark figure*), and the true toll, including fatalities, may well exceed that of alcohol or drug-related incidents (Ref). Assessing sleepiness remains challenging - both subjectively and objectively - even though its evaluation is crucial, as the absence of sleepiness is a legal requirement for safe driving.

Not only does this apply to a driver’s own decision to operate a vehicle, but also to authorities, for example in the context of a sleep-related accident. In traffic medicine and for licensing authorities, assessing excessive daytime sleepiness (EDS) is essential for determining long-term fitness to drive (1). At the same time, enforcement agencies and judicial authorities would benefit from the availability of simple road-side tests for sleepiness after an accident or even before departure. Such testing could be based on behavioral or physical signs of sleepiness (2), or detect metabolic changes in, for example, oral fluid as legally admissible evidence of sleepiness, analogously to current breath-alcohol testing (3, 4).

As part of one such study aimed at characterizing metabolic changes and neurophysiological, subjective and neurobehavioral consequences of insufficient sleep (5), we report the effects of a controlled total 8-hour sleep loss, induced by either cumulative sleep restriction or total sleep deprivation, on measures of simulated driving that have real-world relevance. Using a high-end driving simulator, we comprehensively assessed performance in naturalistic driving and vigilance tests.

While previous studies have shown that both acute sleep deprivation and chronic sleep restriction impair driving performance, a recent systematic review of sleep loss effects on young drivers (6) highlights that these conditions are typically examined in isolation, using heterogeneous sleep-loss protocols and outcome measures. Furthermore, this literature is characterized by substantial variation in study designs, performance metrics, and overall evidence quality, limiting direct comparisons across sleep-loss paradigms. Individual experimental studies further illustrate this heterogeneity, with partial versus total sleep deprivation producing different impairment patterns (7), performance decrements depending on task demands and time on task (8), and outcome-specific effects even within partial restriction paradigms (9). Moreover, many studies rely on small samples and insufficiently control for circadian phase or withdrawal effects, further complicating cross-study comparisons (6).

As a result, it remains unclear whether equivalent amounts of sleep loss incurred acutely versus cumulatively produce comparable impairments across specific driving-relevant performance domains. In the present study, we uniquely combine tightly controlled sleep–wake protocols with a high-fidelity driving simulator and domain-specific performance metrics. This approach allows a direct comparison of cumulative and acute sleep loss under standardized naturalistic conditions.

The purpose of this study was to identify which performance domains are affected under different conditions of equivalent sleep loss and to translate these effects into tangible and relatable messages. This information may help drivers make better-informed decisions about their ability to drive safely.

## Methods

### Study Design

These results presented here were obtained as secondary outcomes from a randomized, controlled, crossover trial that aimed to investigate metabolomics-based biomarkers for sleepiness. The study protocol for this trial was pre-registered and published (5).

Of 20 recruited volunteers (healthy young men, habitually sleeping 7-9 hours per night) 17 fully completed the protocol, and 3 partially completed it. The demographic characteristics of the participants are provided in Table 1. Primary outcomes are reported elsewhere (Scholz et al., submitted.)

**Table 1.**
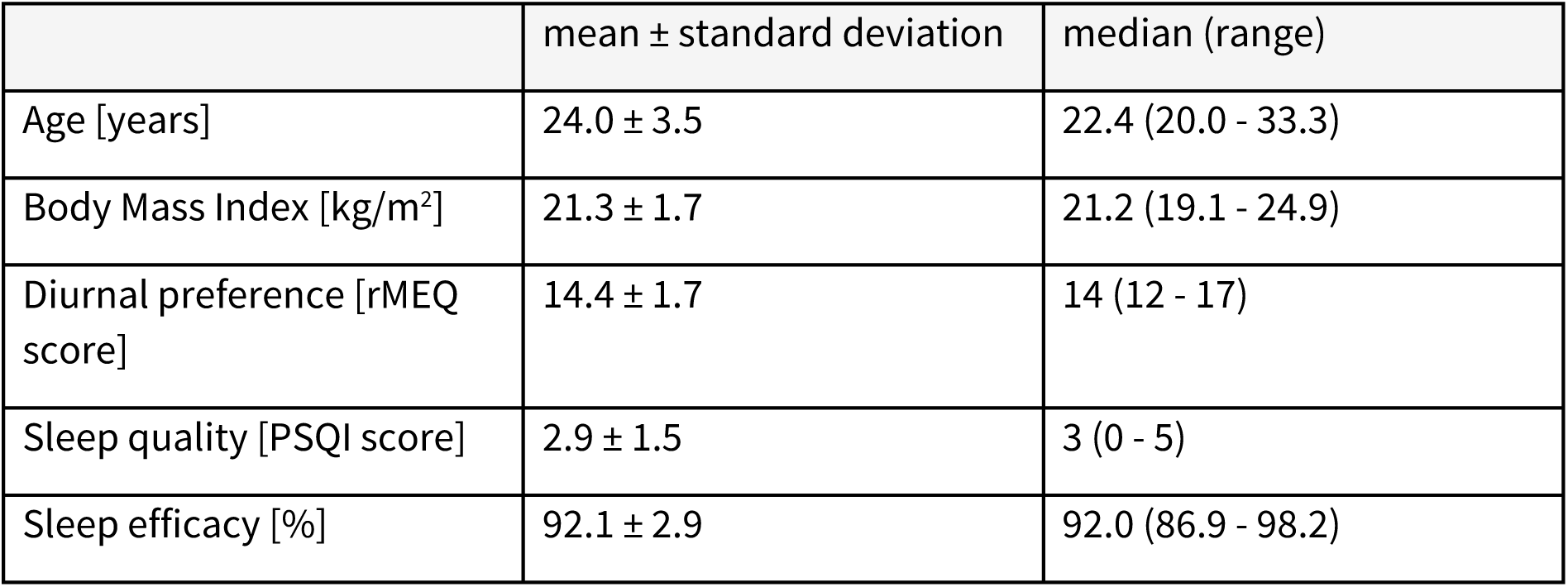
Demographic and sleep-related characteristics of study participants (n = 20). Age is reported at the first day of the first intervention. Diurnal preference was assessed using the reduced Morningness–Eveningness Questionnaire (rMEQ, 30). Subjective sleep quality was assessed using the Pittsburgh Sleep Quality Index (PSQI, (31)). Validated German versions of the rMEQ and PSQI were used. Sleep efficacy was calculated as total sleep time divided by time in bed (8 h) and was determined by polysomnography during the screening night. Values are presented as mean ± standard deviation and median (range).

| | mean ± standard deviation | median (range) |
| --- | --- | --- |
| Age [years] | 24.0 ± 3.5 | 22.4 (20.0 - 33.3) |
| Body Mass Index [kg/m<sup>2</sup>] | 21.3 ± 1.7 | 21.2 (19.1 - 24.9) |
| Diurnal preference [rMEQ score] | 14.4 ± 1.7 | 14 (12 - 17) |
| Sleep quality [PSQI score] | 2.9 ± 1.5 | 3 (0 - 5) |
| Sleep efficacy [%] | 92.1 ± 2.9 | 92.0 (86.9 - 98.2) |

### Driving Simulator

Drive and vigilance data were acquired in a highly immersive, custom-built driving simulator described elsewhere in detail (1). In brief, the simulator is based on a full BMW i3 chassis, with each wheel’s shock absorber replaced by spindle actuators, providing motion cues primarily reflecting surface imperfections, acceleration, and braking. Controls for throttle, brake, indicator, and steering were modified to enable (I) vehicle control and dynamics, (II) integration into the virtual environment displayed in (mirrors, rear-view monitor) and 270° around the car (via five DLP Laser Phosphor projectors) and (III) data acquisition at 120 Hz (SILAB 7.0, WIVW, Würzburg, Germany).

### Procedure, Driving and Vigilance Task

Prior to the simulated driving task, participants were either well-rested (Condition C) or subjected to total sleep deprivation (Condition D), achieved by skipping one entire night (8 h deficit, acute sleep loss), or cumulative sleep restriction (Condition R), achieved by curtailing sleep by two hours each night over four consecutive nights (4 × 2 h deficit, cumulative sleep loss). Both interventions resulted in an equivalent total sleep loss of eight hours. Participants underwent each condition in a randomized, controlled, crossover fashion. Adherence to the protocol and behavioral instructions were closely monitored by the study team in a controlled sleep-laboratory environment.

After self-reporting subjective sleepiness (Karolinska Sleepiness Scale, KSS, version A), participants completed a customized driving task consisting of 9 modules. Driving started with a 12-min monotonous nighttime car-following scenario featuring a sound-based vigilance task (three equal modules 1–3). After a 1.5-min transition (module 4) and a brightly lit winding tunnel (0.5 min, module 5), open driving continued in a 5-min foggy early-morning rural scenario (modules 6-8). After another short transition, a police car approached from behind, overtook (module 9) and subsequently signaled the driver to stop. The total driving time was approximately 22 minutes for 28.5 kilometers track-length. During screening, participants were familiarized with the driving simulator in a drive encompassing Module 1-8 (Condition S).

Modules 1–3 included a tone-based vigilance task: drivers heard a regular sequence of tones (3,400 ms intervals) and were instructed to press a button on the left indicator stalk whenever a tone was omitted (10).

### Data analysis

Driving and vigilance data were analyzed for modules 1-9 according to comparable studies on sleepiness and driving (1, 10) using R (R version 4.2.2, R-Studio 2024 12.0.0) and GraphPad Prism (GraphPad Prism 10.4.1., Dotmatics, Hertfordshire, England). All analyses steps are reported in the supplemental material and R-Markdown.

In brief, collected lateral (lane postion, LP), longitudinal (distance to lead car, d), and speed (s) data were evaluated as lateral, longitudinal and speeding control by calculating the standard deviations of each the lateral position (SDLP), the distance to the lead vehicle (SDD), and speed (SDS). We furthermore evaluated lane keeping ability by way of several measures for inappropriate line crossings (ILC), i.e., total count (tcILC), total duration (ttILC), and average duration per event (atILC). We assessed steering stability by counting steering wheel reversals (cSWR). Vigilance was evaluated by calculating sensitivity (Sens, proportion of correct responses when a signal/omission occurs, in percent) and reaction time (RT, time to react to omission of sound signal, in milliseconds).

In order to compare overall driving performance, i.e., across all variables between conditions, an Integrated Driving Score per drive (IDS) or per module (IDSM) was obtained by summing the z-scores of the respective variables (11) and then normalizing the sum based on its standard deviation. In the summation, any missing or excluded values are treated as 0, i.e., average performance. Here, higher values, expressed in units of standard deviations (sigma, σ), indicate worse driving performance.

### Descriptive statistics, repeated measures ANOVA and correlational analysis

Means and standard errors (SE) for each variable were computed either per drive or per module. Treating condition (c) as a fixed effect and visit and participant as random effects in linear mixed models, we evaluated model statistics (F), statistically significant differences between conditions (*p* < 0.05), and the magnitude of pairwise comparisons.

Effect sizes were calculated as Cohen’s *d* and interpreted as negligible, small, medium, or large (for details, see supplemental information). For each outcome, an omnibus linear mixed-effects model was evaluated first. Pairwise comparisons between conditions were Tukey-adjusted within each outcome. To control for multiple testing across outcomes addressing the same domain construct, p-values were additionally adjusted within predefined performance domains using the Holm–Bonferroni procedure (lateral control: five outcomes; vigilance: two outcomes). Domain-level Holm–Bonferroni correction was applied separately for each pairwise contrast across outcomes within the same domain.

Subjective sleepiness (KSS) and the Integrated Driving Score (IDS) were analyzed as single outcomes and were not adjusted across domains. F or descriptive synthesis, outcomes were assigned to grouping categories based on the pattern and statistical significance of pairwise comparisons between normal sleep (C), cumulative sleep restriction (R), and acute sleep deprivation (D), as summarized in Tables 2 and 3. Grouping categories were defined as follows: Group 3, significant stepwise differences between all conditions (C < R < D); Group 2, no difference between C and R but significantly higher values in D (C = R < D); Group 1, no difference between R and D but significantly higher values than C (C < R = D); and Group 0, no significant differences between conditions (C = R = D).

**Table 2.** Effects of sleep condition on subjective sleepiness, driving performance, and vigilance outcomes. Linear mixed-effects model results are shown for subjective sleepiness, overall driving performance, and operational and vigilance outcomes across normal sleep (C), cumulative sleep restriction (R), and acute sleep deprivation (D). For each outcome, the omnibus model test (F-statistic), estimated pairwise contrasts (estimate ± standard error), and corresponding p-values are reported. Pairwise p-values are Tukey-adjusted within outcome; domain-level correction (Holm–Bonferroni) was applied across outcomes within the lateral control and vigilance domains. Effect sizes are reported as Cohen’s *d* with conventional qualitative interpretation. The final column provides a cross-reference to outcome groupings summarized in Table 3.

| Domain | Variable | Contrast | Estimate<br>(SE) | F_or_t | p<br>(LMM) | p<br>(Tukey) | Sign.<br>(Tukey) | p<br>(Holm) | Sign.<br>(Holm) | Cohen's d | Interpretation | Group<br>(Table 3) |
| --- | --- | --- | --- | --- | --- | --- | --- | --- | --- | --- | --- | --- |
| subjective sleepiness | KSS27 |  |  | 27.74 | 0.0000 |  | **** |  |  |  |  |  |
|  |  | C - D | -2.94 (0.39) | -7.36 |  | 0.0000 | **** |  |  | -2.2049 | large |  |
|  |  | C - R | -1.41 (0.40) | -3.46 |  | 0.0044 | ** |  |  | -1.0562 | large | 3 |
|  |  | D - R | 1.52 (0.39) | 3.83 |  | 0.0017 | ** |  |  | 1.0474 | large |  |
| overall performance | IDS |  |  | 15.17 | 0.0000 |  | **** |  |  |  |  |  |
|  |  | C - D | -0.98 (0.19) | -5.26 |  | 0.0000 | **** |  |  | -1.0316 | large |  |
|  |  | C - R | -0.24 (0.18) | -1.28 |  | 0.4161 | ns |  |  | -0.4533 | small | 2 |
|  |  | D - R | 0.74 (0.19) | 3.96 |  | 0.0012 | ** |  |  | 0.6601 | medium |  |
| lateral control | SDLP |  |  | 22.17 | 0.0000 |  | **** |  |  |  |  |  |
|  |  | C - D | -4.61 (0.71) | -6.48 |  | 0.0000 | **** | 0.0000 | **** | -1.1307 | large |  |
|  |  | C - R | -1.34 (0.71) | -1.87 |  | 0.1642 | ns | 0.8208 | ns | -0.4028 | small | 2 |
|  |  | D - R | 3.27 (0.72) | 4.52 |  | 0.0003 | *** | 0.0013 | * | 0.753 | medium |  |
|  | atILC |  |  | 9.08 | 0.0008 |  | *** |  |  |  |  |  |
|  |  | C - D | -0.91 (0.23) | -3.98 |  | 0.0011 | ** | 0.0045 | ** | -1.1529 | large |  |
|  |  | C - R | -0.18 (0.23) | -0.76 |  | 0.7276 | ns | 1.0000 | ns | -0.5427 | medium | 2 |
|  |  | D - R | 0.73 (0.23) | 3.20 |  | 0.0088 | ** | 0.0263 | * | 0.8596 | large |  |
|  | tcILC |  |  | 7.19 | 0.0028 |  | ** |  |  |  |  |  |
|  |  | C - D | -0.98 (0.29) | -3.48 |  | 0.0043 | ** | 0.0086 | ** | -0.9444 | large |  |
|  |  | C - R | -0.14 (0.29) | -0.49 |  | 0.8784 | ns | 1.0000 | ns | -0.4991 | small | 2 |
|  |  | D - R | 0.84 (0.29) | 2.98 |  | 0.0151 | * | 0.0301 | * | 0.7557 | medium |  |
|  | cSWR |  |  | 6.90 | 0.0034 |  | ** |  |  |  |  |  |
|  |  | C - D | -14.72 (3.99) | -3.69 |  | 0.0025 | ** | 0.0074 | ** | -0.8668 | large |  |
|  |  | C - R | -7.16 (4.07) | -1.76 |  | 0.1993 | ns | 0.8208 | ns | -0.5944 | medium | 1 |
|  |  | D - R | 7.56 (3.99) | 1.89 |  | 0.1582 | ns | 0.3164 | ns | 0.3775 | small |  |
|  | ttILC |  |  | 2.65 | 0.0863 |  | ns |  |  |  |  |  |
|  |  | C - D | -4.77 (2.38) | -2.00 |  | 0.1287 | ns | 0.1287 | ns | -0.5691 | medium |  |
|  |  | C - R | -0.14 (2.43) | -0.06 |  | 0.9982 | ns | 1.0000 | ns | -0.5427 | medium | 0 |
|  |  | D - R | 4.63 (2.38) | 1.94 |  | 0.1435 | ns | 0.2869 | ns | 0.5475 | medium |  |
| longitudinal control | SDD |  |  | 5.49 | 0.0094 |  | ** |  |  |  |  |  |
|  |  | C - D | -3.84 (1.34) | -2.87 |  | 0.0200 | * |  |  | -0.6799 | medium |  |
|  |  | C - R | -0.03 (1.33) | -0.03 |  | 0.9996 | ns |  |  | -0.1057 | negligible | 2 |
|  |  | D - R | 3.81(1.33) | 2.87 |  | 0.0200 | * |  |  | 0.6587 | medium |  |
| speed control | SDS |  |  | 1.89 | 0.1686 |  | ns |  |  |  |  |  |
|  |  | C - D | -0.38 (0.20) | -1.90 |  | 0.1564 | ns |  |  | -0.5937 | medium |  |
|  |  | C - R | -0.23 (0.11) | -1.17 |  | 0.4795 | ns |  |  | -0.196 | negligible | 0 |
|  |  | D - R | 0.15 (0.20) | 0.77 |  | 0.7252 | ns |  |  | 0.2334 | small |  |
| vigilance | RT |  |  | 1.55 | 0.2295 |  | ns |  |  |  |  |  |
|  |  | C - D | 12.92(26.40) | 0.49 |  | 0.8768 | ns | 0.8768 | ns | 0.0712 | negligible |  |
|  |  | C - R | -31.85 (26.90) | -1.18 |  | 0.4712 | ns | 0.9492 | ns | -0.1818 | negligible | 0 |
|  |  | D - R | -44.77(26.40) | -1.70 |  | 0.2232 | ns | 0.4464 | ns | -0.2699 | small |  |
|  | Sens |  |  | 2.47 | 0.1005 |  | ns |  |  |  |  |  |
|  |  | C - D | 4.50 (2.14) | 2.10 |  | 0.1057 | ns | 0.2114 | ns | 0.2547 | small |  |
|  |  | C - R | 1.07 (2.19) | 0.49 |  | 0.8769 | ns | 0.9492 | ns | 0.0624 | negligible | 0 |
|  |  | D - R | -3.43(2.14) | -1.60 |  | 0.2592 | ns | 0.4464 | ns | -0.1774 | negligible |  |

**Table 3.**
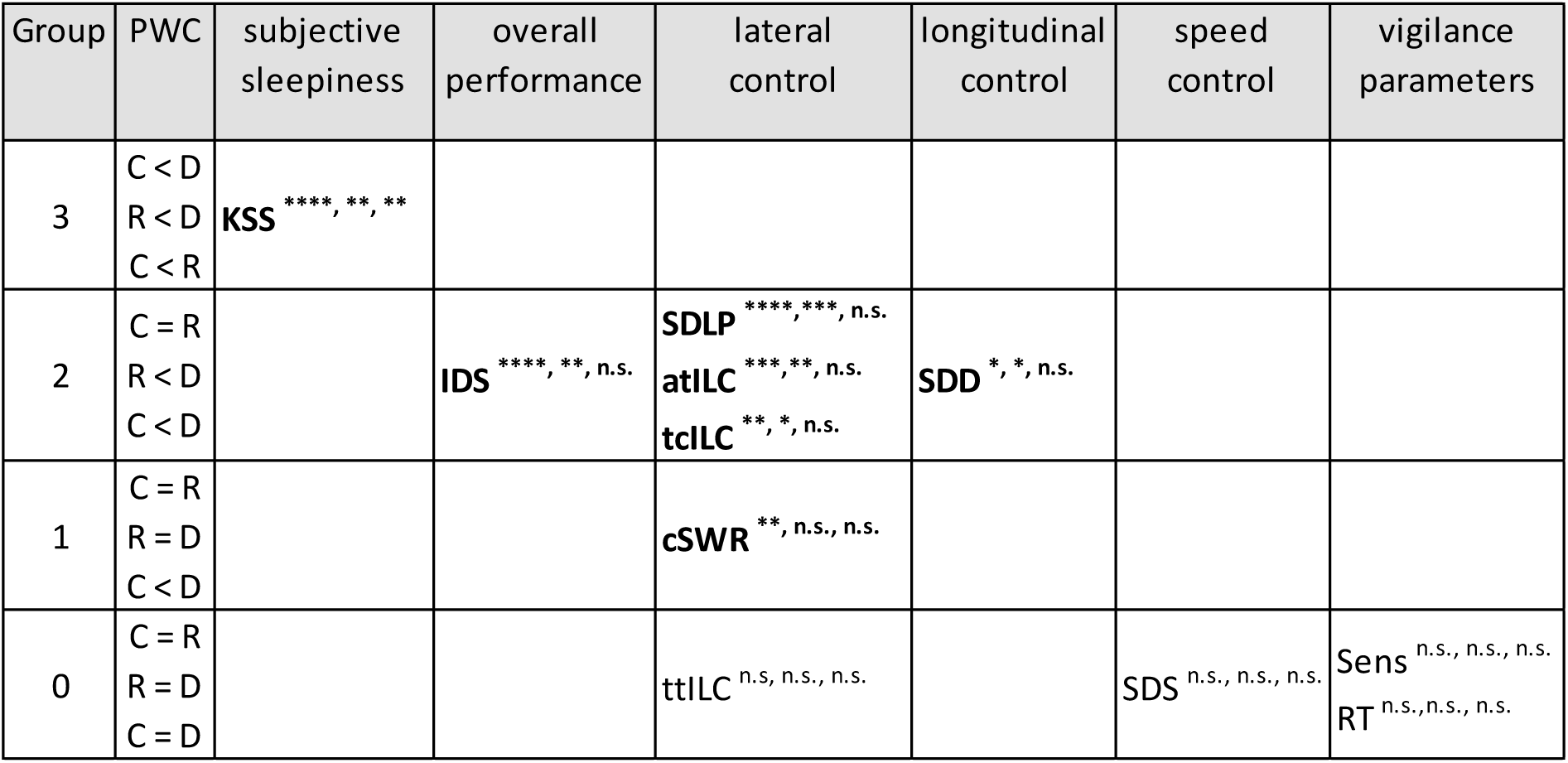
Summary of relative drive-level effects of sleep interventions across outcome domains. Outcomes are grouped according to the pattern of significant pairwise comparisons (PWC) between normal sleep (C), cumulative sleep restriction (R; 8 h total sleep loss), and acute sleep deprivation (D; 8 h total sleep loss). Significant and non-significant differences between condition means are indicated by “<” and “=”, respectively. Groups are defined stepwise based on the presence of significant differences across all three pairwise comparisons, ranging from Group 3 (significant differences for all pairs: C < D, R < D, C < R) to Group 0 (no significant differences: C = D, R = D, C = R). Outcomes are organized by functional domain (subjective sleepiness, overall performance, lateral control, longitudinal control, speed control, and vigilance). The order of symbols reflects the sequence of pairwise comparisons. Bold type indicates outcomes showing a significant drive-level correlation with subjective sleepiness (KSS). Statistical details and correlation analyses are provided in the Supplemental Information.

Time-on-task (t.o.t.) effects were obtained for changes of variables recorded in equal modules 1-3 based on repeated measures analysis of variance (ANOVA). Significance of each fixed term condition (c) or module (m) as well as of any interaction between condition and module (c * m) evaluated differences of time-on-task effects between conditions. Module was modelled both as a categorical factor and as a linear trend to distinguish overall time-on-task effects from directional changes across modules. Planned within-module contrasts between normal sleep (C) and acute sleep deprivation (D) were specified a priori and are reported irrespective of omnibus interaction tests. Results of t.o.t.-analyses are reported hierarchically, with primary inferences based on repeated-measures analyses of condition, module, and their interaction. Significant interactions were clarified using a priori planned pairwise contrasts within identical modules, while descriptive figures illustrate the underlying trajectories. Within-condition analyses were conducted for completeness but are not reported in detail, as they are subsumed by the interaction and planned contrasts. Correlation to subjective sleepiness (KSS) before the drive was evaluated for drive-, module- and t.o.t.-data (for details, see supplemental information).

## Results

### Subjective sleepiness before the drive

Mean subjective sleepiness (KSS) increased across sleep conditions, with higher ratings observed under cumulative (R) and acute (D) sleep loss compared to normal sleep (C), indicating progressively elevated perceived sleepiness prior to the naturalistic driving and vigilance task (Figure 1 a, Table 2). Significant main effects of sleep condition were observed (F = 27.74, *p* < 0.0001), and pairwise comparisons demonstrated a near-stepwise increase in KSS across conditions (C < R < D, group 3, see Table 3), with higher scores in R compared to C (C–R: −1.41 ± 0.41, *p* = 0.0044) and in D compared to both C (C–D: −2.94 ± 0.40, *p* < 0.0001) and R (D–R: 1.53 ± 0.40, *p* = 0.0017). Notably, despite identical total sleep loss, acute sleep deprivation was associated with significantly higher subjective sleepiness than cumulative sleep restriction.

**Figure 1.**
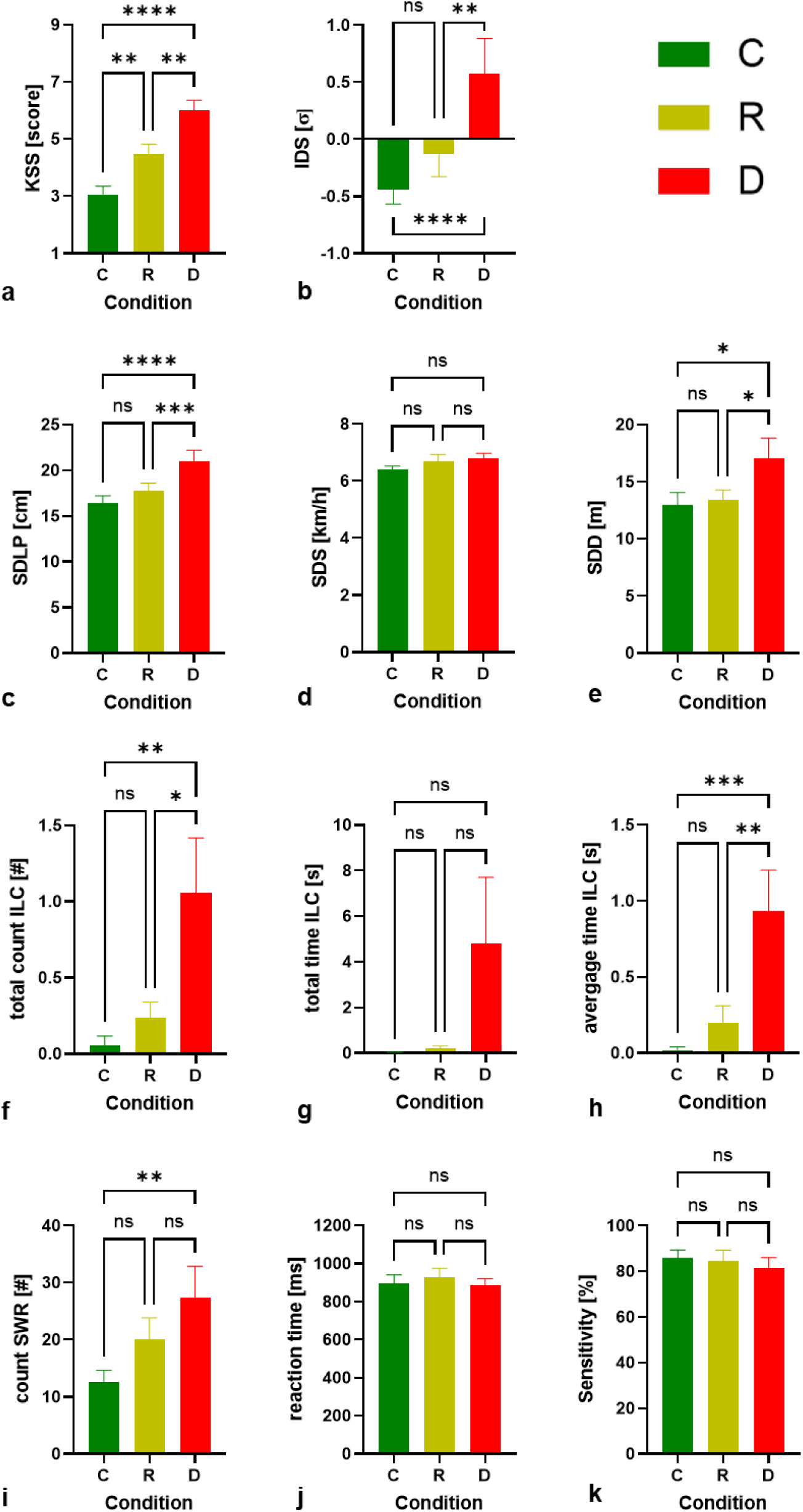
Drive-level comparison of subjective sleepiness, driving and vigilance outcomes across sleep conditions. Panels (a–k) show mean values for normal sleep (C, control), cumulative sleep restriction (R; 4 nights with 2 h sleep reduction per night), and acute sleep deprivation (D; 24 h continuous wakefulness). Outcomes include subjective sleepiness assessed by the Karolinska Sleepiness Scale (KSS; a), overall driving performance quantified by the Integrated Driving Score (IDS; b), operational driving measures (c–i), and vigilance measures (j, k). Bars represent means ± SEM. Statistically significant pairwise differences between conditions are indicated in brackets (p < 0.05, corrected for multiple comparisons). Colors are used for visualization only; conditions are additionally distinguished by bar position and labeling, and figures remain interpretable in grayscale.

### Overall performance by IDS

Mean Integrated Driving Scores (IDS) increased from C to R to D, consistent with poorer overall performance across the assessed variables and visually suggesting a stepwise pattern (Figure 1 b, Table 2). Significant differences in IDS were observed under conditions of acute sleep loss (F = 15.17, *p* < 0.0001), with higher scores in D compared to both C (C–D: −0.98 ± 0.19, *p* < 0.0001) and R (D–R: 0.74 ± 0.19, *p* = 0.0012). In contrast to the pattern observed for subjective sleepiness, and despite similar total sleep loss, cumulative sleep loss did not result in a statistically significant difference in IDS compared with normal sleep (C–R: p = 0.4162, corresponding to Group 2; see Table 2 and 3).

### Descriptive analysis of drive-level outcomes

We then analyzed the effects of acute and cumulative sleep loss on each driving and vigilance parameter across drives and found that lateral and longitudinal control outcomes exhibited significant differences relative to normal sleep, whereas no significant effects were observed for speed-related or vigilance outcomes (see Table 3 for an overview).

Among the lateral control outcomes, three measures exhibited a response pattern comparable to the Integrated Driving Score (IDS) following a total of 8 h of sleep loss, with significant differences observed only under acute sleep loss (D) relative to both normal sleep (C) and cumulative sleep loss (R), and no significant difference between C and R (Group 2). The standard deviation of lateral position (SDLP, Figure 1 c) showed a Group 2 response pattern with pronounced mean increases and statistically significant differences under acute sleep loss (D) relative to C and R. In contrast, the average time and total count of inappropriate line crossings (atILC and tcILC) followed the same response pattern as SDLP and showed substantial relative increases in mean values under acute sleep loss (Figure 1 h, f), accompanied by large effect sizes. However, greater inter-individual variability in these measures limited statistical significance in comparisons with normal sleep (C) and cumulative sleep loss (R). Along these lines, the mean count of steering wheel reversals (cSWR), defined as abrupt steering movements in both directions exceeding a total of 6° within a 1-s time window, showed a near-linear increase in mean values across conditions C, R, and D with moderate variability (Figure 1 i) and exhibited a Group 1 response pattern, with a statistically significant difference between acute sleep loss and normal sleep but no difference between cumulative and acute sleep loss (Table 2 and 3).

The ability to control the distance to the lead vehicle was assessed using the standard deviation of this distance (SDD, Figure 1 e). Mean SDD values showed little change between normal sleep and cumulative sleep loss but increased under acute sleep loss, exhibiting a Group 2 response pattern with statistically significant differences relative to normal sleep and medium effect sizes. It should be noted that SDD was derived from relatively short observation periods (Modules 1–3).

In contrast, neither speed control (SDS, Figure 1 d). total time outside the lane (ttILC, Figure 1 g) nor any of the measures obtained from the tone-based vigilance task (reaction time or sensitivity, Figure 1 j and k, assessed in Modules 1–3) showed significant differences across sleep conditions, corresponding to a Group 0 pattern.

### Module-level descriptive analysis and time-on-task effects

The full drives consisted of nine modules which, apart from the identical Modules 1–3, differed in length and driving task characteristics. To compare driving performance between condition (c) and module (m), a module-wise Integrated Driving Score (IDSM) was calculated based on the outcomes available in all modules, excluding distance and vigilance measures that were only obtained in Modules 1–3 (full module data, see Supplementary Materials).

As shown in Figure 2 a, acute sleep loss was associated with consistently higher mean IDSM values across modules, with particularly pronounced increases emerging after the first module. Repeated-measures analyses restricted to the identical Modules 1–3 revealed a significant Condition × Module interaction when modelling module as a linear trend (F(2,131) = 3.32, p = 0.039, Table 4), indicating differential time-on-task effects between sleep conditions. Planned within-module contrasts between normal sleep and acute sleep deprivation showed no significant difference in Module 1, but statistically significant differences in Modules 2 (p = 0.004) and 3 (p = 0.009), both associated with large effect sizes (|d| = 0.92 and 0.88, respectively, see Table 5). Consistent with this interaction, IDSM increased significantly across modules within the sleep-deprivation condition alone (Table 4), whereas no such effects were present under normal sleep or cumulative sleep loss.

**Figure 2.**
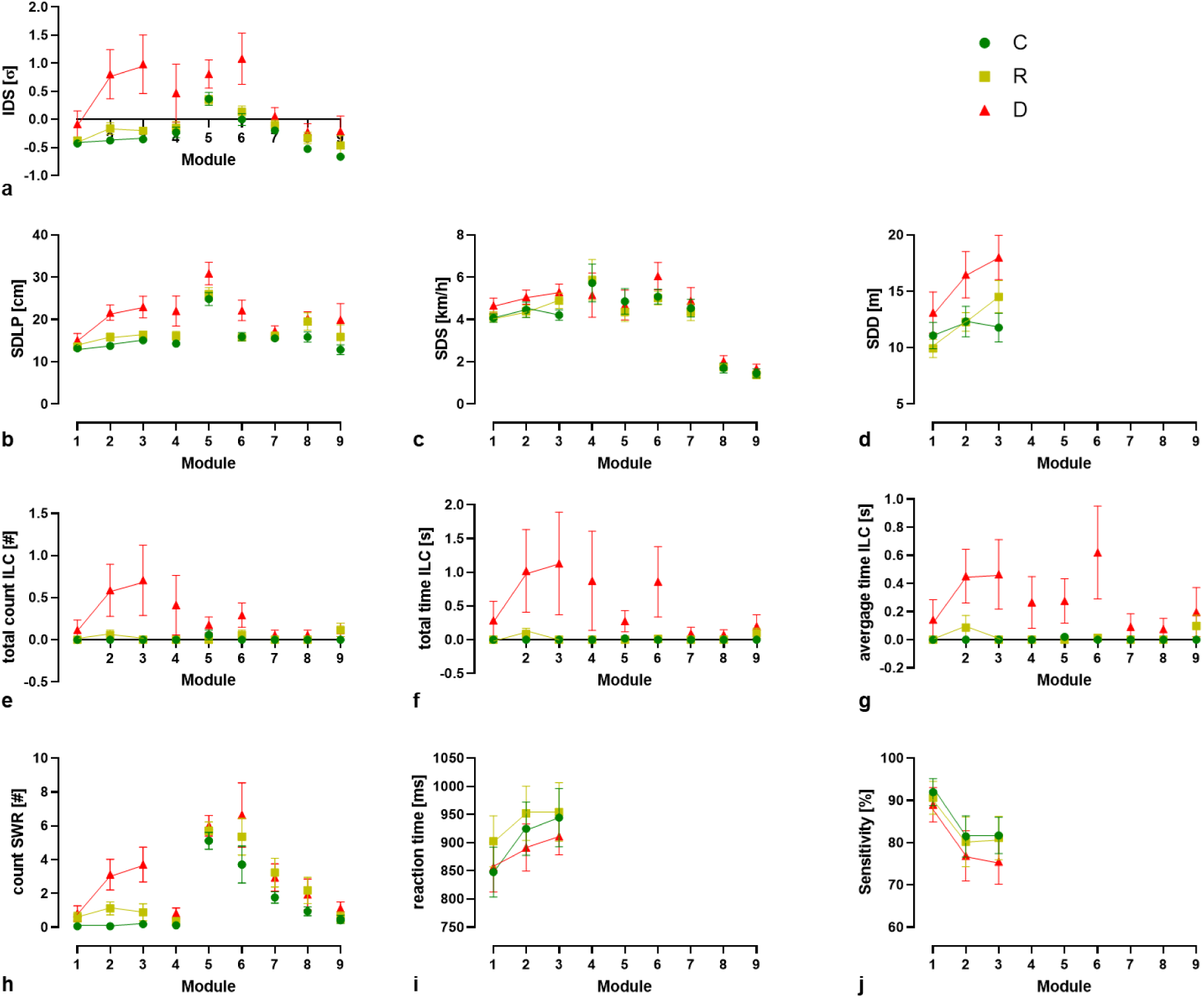
Module-level development of driving and vigilance outcomes across sleep conditions. Panels (a–j) show mean values across successive driving modules for normal sleep (C, control), cumulative sleep restriction (R; 4 nights with 2 h sleep reduction per night), and acute sleep deprivation (D; 24 h continuous wakefulness). Outcomes are ordered to match Figure 1 and include integrated driving performance (a), operational driving measures (d–h), and vigilance measures (i, j). Symbols represent condition-specific means ± SEM. Lines connecting module-specific values indicate repeated measurements within identical driving tasks and serve to illustrate time-on-task development; statistical effects of condition and time-on-task were assessed using linear mixed-effects models and are reported in the Results. No statistical annotations are shown in the figure. Colors are used for visualization only; conditions are additionally distinguished by symbol shape and remain interpretable in grayscale.

**Table 4.**
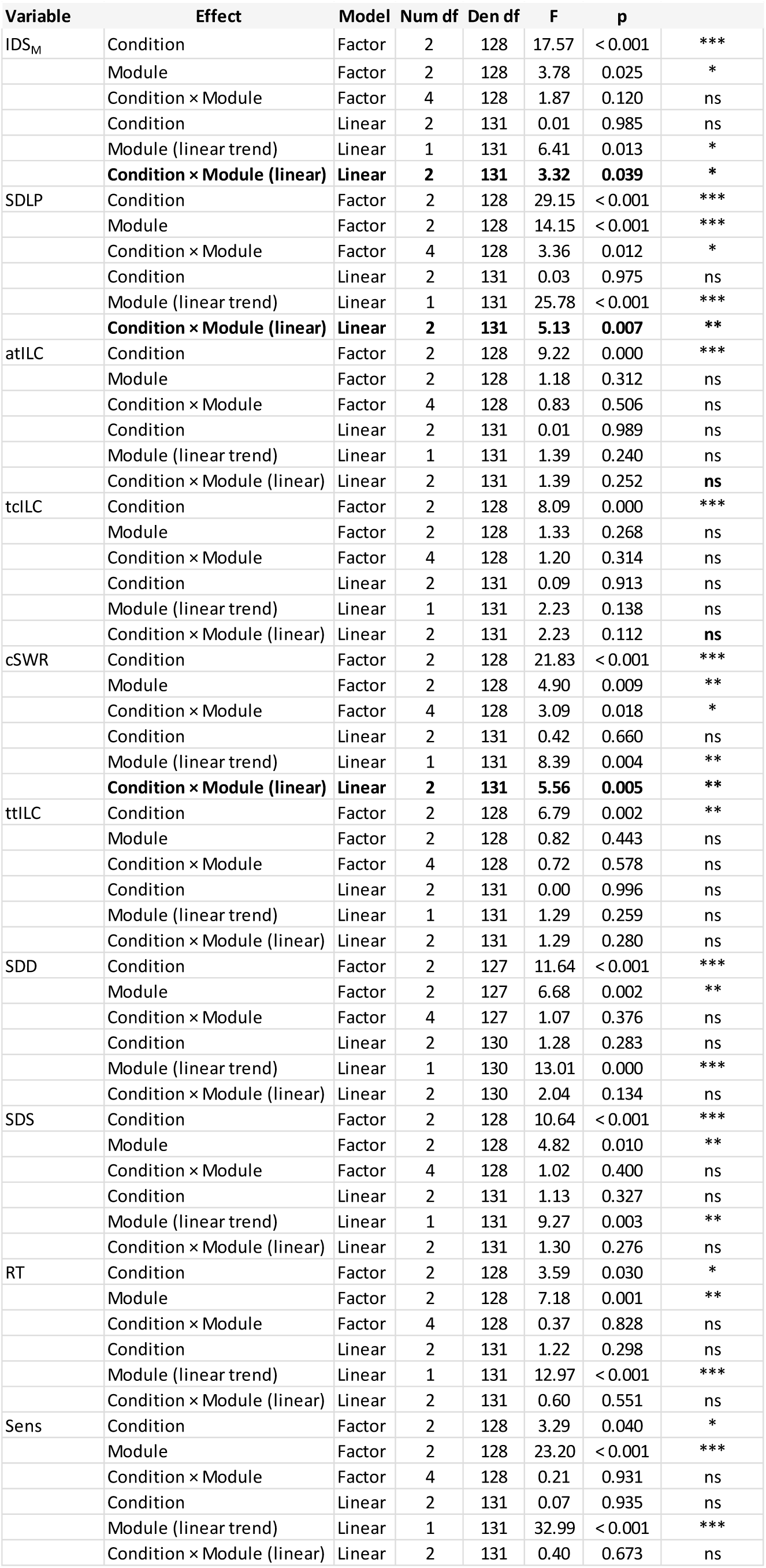
Omnibus linear mixed-effects model tests of sleep condition and time-on-task effects on module-level outcomes. For each outcome variable, omnibus linear mixed-effects model tests are reported for fixed effects of sleep condition and time-on-task (module). Time-on-task was modeled both as a categorical factor (to test non-linear module-specific effects) and as a numeric variable (to test linear time-on-task trends). The table lists numerator and denominator degrees of freedom, F-statistics, and corresponding p-values for each tested effect. Models included participant as a random intercept. Planned pairwise comparisons between normal sleep and acute sleep deprivation (C–D) are reported separately in Table 5.

**Table 5.**
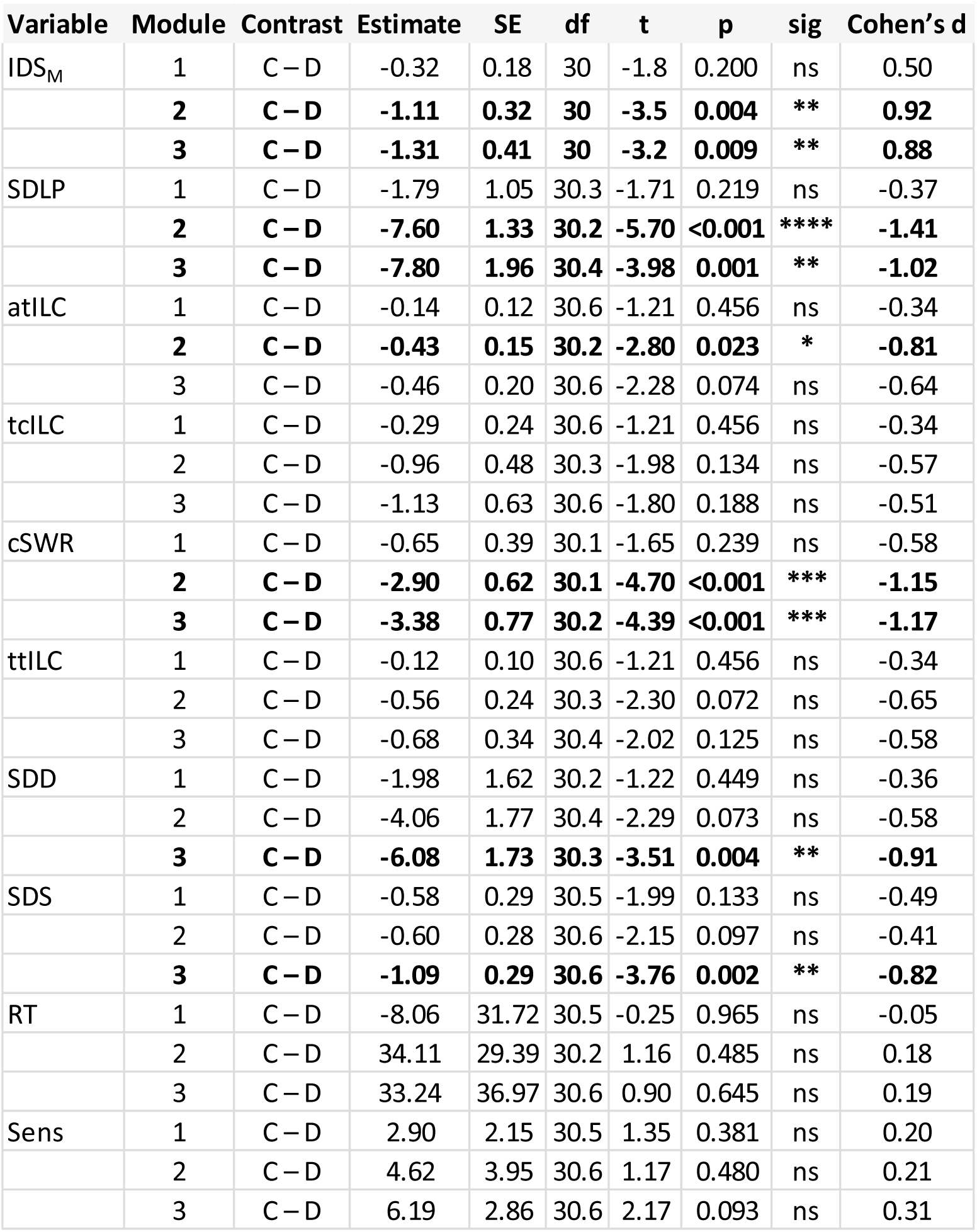
Planned pairwise comparisons between normal sleep and acute sleep deprivation across driving modules 1-3. This table reports planned pairwise contrasts between normal sleep (C) and acute sleep deprivation (D) for each outcome variable at the module level 1-3. For each contrast, the estimated difference (estimate ± standard error), degrees of freedom, t-statistic, p-value, and corresponding effect size (Cohen’s *d*) are shown. Planned comparisons were specified *a priori* and evaluated within the linear mixed-effects modeling framework. Significance codes are provided for descriptive purposes.

When the operational driving parameters comprising the IDSM were examined individually, only the lateral control outcomes SDLP (swerving) and cSWR (abrupt steering corrections) showed statistically significant time-on-task effects, as confirmed by repeated-measures ANOVA.

In detail, SDLP showed significant main effects of Condition and Module (see Table 4), as well as a significant Condition × Module interaction when modelling module as a linear trend (F(2,131) = 5.13, p = 0.007), indicating condition-dependent time-on-task effects.

Specifically, the time-on-task effect was significantly greater following acute sleep loss than after normal sleep or cumulative sleep loss. Planned within-module contrasts between normal sleep and acute sleep deprivation were not significant in Module 1, but were highly significant in Modules 2 (p <0.001 and 3 (p = 0.001), both associated with large effect sizes (d = 1.41 and d = 1.02, respectively, Table 5). In absolute terms, mean SDLP increased by 7.9 cm (+53%) from Module 1 to Module 3 following acute sleep loss, with most of the increase occurring within the first 4–6 minutes and continuing across the 12-minute monotonous night-time drive.

Similarly, the frequency of abrupt steering wheel reversals (cSWR) exhibited significant main effects of Condition and Module, as well as a significant Condition × Module interaction for the linear time-on-task trend (F(2,131) = 5.56, p = 0.005, Table 4). As for SDLP, the time-on-task effect was significantly greater following acute sleep loss than under normal sleep or cumulative sleep loss. Planned contrasts between normal sleep and acute sleep deprivation were significant in Module 2 and3 (both p <0.001), both showing large effect sizes (d = 1.15 and d = 1.17, respectively, Table 5).

Apart from SDLP and cSWR, the remaining operational driving parameters did not show statistically significant condition-dependent linear time-on-task effects. While several measures exhibited significant main effects of condition and/or module, no additional Condition × Module interactions reached significance when modelling module as a linear trend. For the vigilance measures reaction time and sensitivity, strong time-on-task effects were observed, reflected in pronounced linear module trends; however, these trends were not significantly modulated by sleep condition (Table 4).

Taken together, and as illustrated in Figure 2, similar differential time-on-task effects were apparent across all driving performance measures following acute sleep loss (Figure 2 b-h), whereas the vigilance outcomes showed largely parallel trajectories and ranges across conditions (Figure 2 i, j) , indicating qualitatively different time-on-task patterns between operational and vigilance domains.

### Correlation to subjective sleepiness

To examine associations between subjective sleepiness and performance, Karolinska Sleepiness Scale (KSS) scores were correlated with drive- and module-level performance measures. At the overall drive level, higher KSS scores were significantly associated with poorer overall driving performance, as reflected by the integrated driving score (IDS, p = 0.0006). In addition, higher subjective sleepiness was associated with increased impairment in several operational outcomes, primarily lateral control measures, including SDLP, total and average ILC, and cSWR (all p < 0.001), as well as with speed and distance control (SDD, p = 0.01). In contrast, no significant associations were observed for SDS, ttILC, or for the vigilance outcomes sensitivity and reaction time.

When module-level data were pooled across sleep conditions, KSS remained significantly associated with lateral control measures SDLP (KSS × Module, p = 0.029) and cSWR (p = 0.013). While trend-level associations with speed and distance control (SDS: p = 0.053; SDD: p = 0.052) with increasing time-on-task remained, no significant associations were observed for vigilance measures (reaction time, sensitivity), nor were such relationships present when each sleep condition was examined separately.

## Discussion

This study demonstrates that different forms of sleep loss exert distinct effects on subjective sleepiness and on selected aspects of driving-related performance under controlled, naturalistic driving conditions.

Our controlled sleep-wake protocol produced marked and stepwise differences in self-reported sleepiness (KSS) between sufficient sleep, continued sleep restriction, and total sleep deprivation. These findings indicate that a gradually accumulated 8-hour sleep loss through sleep restriction is perceived as less severe than an acute 8-hour loss induced by total sleep deprivation.

This dose-dependent effect of sleep restriction and deprivation on subjective sleepiness is well in line with available studies (12–14). Interestingly, some studies report significant increases in subjective sleepiness already after a single night of sleep restriction (13) , whereas performance effects such as sustained attention or reaction time were less consistently affected than after total deprivation. For example, a meta-analysis of 44 studies by Wüst et al. (14) found a robust effect on subjective sleepiness already after one night of sleep restriction to 2-6 h sleep opportunity, although subjective sleepiness was not associated with sleep duration. While sustained attention was robustly impaired, choice reaction time, cognitive throughput, working memory, or inhibitory control were not affected (14).

Driving, however, places simultaneous demands on multiple cognitive and operational-level processes, requiring the continuous integration of motor execution with sustained attention, visuomotor processing, and rapid decision-making. Against this background, we first assessed overall driving performance at the level of the complete drive using the Integrated Driving Score (IDS), a composite measure that combines indices of operational vehicle control with vigilance-related outcomes such as reaction time and sensitivity.

These vigilance components reflect core cognitive functions, including sustained attention and processing speed, which are directly engaged during prolonged driving.

Visually, IDS increased from normal sleep to sleep restriction and total sleep deprivation, suggesting a sleep-loss–related decline in overall driving performance. However, statistical significance was observed only following total sleep deprivation, indicating that the smaller effects observed after sleep restriction across individual outcomes did not sum to a detectable overall performance deficit, despite an equivalent total sleep loss of 8 hours. Notably, although total sleep loss was matched between conditions, only total sleep deprivation involved a complete absence of sleep, underscoring the potential protective role of even restricted sleep opportunities for maintaining driving performance. This pattern suggests that partial sleep obtained during the restriction nights may have supported compensatory cognitive or operational strategies during driving, mitigating performance impairments that became evident only after a full night without sleep.

Examining the individual outcome measures therefore allows further insight into whether such compensation primarily affects operational or cognitive performance domains. In line with this interpretation, a similar pattern - significant differences observed only following total sleep deprivation - was evident for most lateral control measures, including SDLP, atILC, and tcILC, as well as for the longitudinal control outcome SDD (Figure 1 and Table 3). Qualitatively, this interpretation is further supported by the pattern observed for the number of abrupt steering maneuvers (cSWR), which showed pronounced changes only following total sleep deprivation.

Taken together, these findings indicate a clear impairment of operational vehicle control after acute sleep loss, while suggesting that the cumulative 8-hour sleep deficit distributed across multiple nights was largely compensated, likely due to the continued opportunity for sleep during the restriction period. Even when cumulative sleep loss is matched, prior controlled protocols show that distributed sleep restriction and total sleep deprivation can yield different impairment trajectories, with restricted sleep opportunities altering the time course and apparent severity of neurobehavioral deficits (15–17). In particular, dose–response laboratory studies demonstrate that performance vulnerability depends on both total sleep loss and its temporal distribution (17), supporting the notion that continued sleep opportunity can attenuate or delay detectable impairment on complex tasks.

Importantly, the pattern of preserved overall driving performance occurred despite pronounced and graded increases in subjective sleepiness. Taken together, although KSS scores showed large and statistically significant differences across all pairwise comparisons (p ≤ 0.004, d ≥ 1.05), indicating robust and stepwise intervention effects on subjective sleepiness, the behavioral consequences of a cumulative 8-hour sleep deficit appeared largely attenuated at the level of driving performance. This dissociation suggests that increased subjective sleepiness under sleep restriction was, at least in part, compensated during driving.

Such dissociations between subjective sleepiness and objective performance have been described previously in controlled sleep-restriction and deprivation studies (14, 15, 18, 19). These studies demonstrate that subjective sleepiness can increase steeply and early during sleep restriction, whereas performance impairments may emerge more gradually, depend on task characteristics, or remain limited in complex, engaging tasks, underscoring that subjective state alone is not a sufficient predictor of functional impairment.

Interestingly, neither speed control nor any of the vigilance-related outcomes exhibited significant intervention-related changes. This pattern points to differential effects of sleep loss across performance domains, suggesting that acute 8-hour total sleep deprivation—but not cumulative sleep restriction—is sufficient to induce measurable impairments in operational driving control, while cognitive vigilance measures remain comparatively unaffected under the present experimental conditions.

Such differential sensitivity across performance domains has been described previously, with vigilance impairments being particularly robust in highly controlled laboratory paradigms involving sustained, monotonous attention tasks, such as the psychomotor vigilance task(20). However, the expression of such impairments depends strongly on task characteristics, including duration, complexity, and ecological engagement (21). Meta-analytic and integrative reviews further emphasize that complex, ecologically engaging activities can permit transient compensatory allocation of effort, even as underlying sleep pressure increases (20, 22). In operationally demanding or motivating contexts, such as simulated driving, vigilance measures may therefore be less sensitive to moderate sleep restriction, allowing impairments to manifest more readily in operational control than in vigilance outcomes.

This operational–vigilance dissociation was further examined by analyzing time-on-task effects across the driving modules. Following acute sleep loss, significant module-wise changes were observed for SDLP and cSWR, whereas no significant module effects emerged for any outcome after normal sleep or cumulative sleep restriction. Beyond these statistically significant effects, visual inspection revealed consistent trends toward impairment across modules for all operational outcomes, as well as for the module-wise Integrated Driving Score excluding vigilance measures, specifically after acute sleep deprivation. In contrast, reaction time and sensitivity showed nearly identical starting values and parallel trajectories across modules 1–3, irrespective of the preceding sleep condition. Thus, while operational driving control exhibited progressive degradation over time following acute sleep loss, vigilance performance evolved in a similar manner across all conditions. These patterns suggest a qualitative time-on-task effect that is expressed primarily in operational driving measures under acute sleep deprivation, rather than reflecting a generalized vigilance decline.

To determine whether the operational–vigilance dissociation extended to the relationship between performance and subjective sleepiness, we examined correlations between KSS and the driving outcomes. Given the robust intervention-related effects on subjective sleepiness and its established association with perceived cognitive readiness and vigilance performance (12, 15, 23), this analysis tested whether subjective sleepiness tracked operational and vigilance performance to a similar extent.

At the drive level, higher KSS scores were significantly associated with impairments in the Integrated Driving Score and in individual operational outcomes, including SDLP, SDD, tcILC, atILC, and cSWR, but not with speed control (SDS), ttILC, sensitivity, or reaction time. Similarly, when module-level data were pooled across all conditions, KSS remained a significant predictor of operational impairments over time in SDLP and cSWR, and a marginal predictor for SDS and SDD. In contrast, no such relationships were observed for the vigilance measures (reaction time and sensitivity) with increasing time-on-task, nor did these associations emerge when sleep conditions were examined separately.

Taken together, these relationships extend the operational–vigilance dissociation by further supporting the notion that subjective sleepiness preferentially reflects impairments in operational driving control rather than vigilance performance, as assessed here.

In line with this pattern, prior research indicates that fatigued or sleep-deprived individuals do not always accurately perceive or acknowledge the full extent of their performance impairment, particularly in complex or operational tasks (22, 24). Dorrian et al. (25) for example, showed that self-rated sleepiness and performance do not reliably track objective decrements across tasks and states of wakefulness, with individuals both over- and underestimating their impairments depending on task demands. Similarly, subjective sleepiness or fatigue ratings do not consistently reflect objective decrements in driving performance, underscoring that the relationship between subjective state and functional impairment is domain- and task-dependent (26).

However, it remains to be determined in future studies whether the observed dissociation between increased subjective sleepiness, preserved vigilance performance, and impaired driving control reflects a meaningful mismatch that may contribute to drivers’ overestimation of their driving ability after sleep loss. Alternatively, this apparent discrepancy may be attributable to methodological factors, such as limited sensitivity or duration of the vigilance task, which could have reduced the likelihood of detecting subtle cognitive impairments. deficits.

## Conclusions Limitations & Outlook

In total, the present study underscores that a single night of total sleep deprivation is sufficient to produce substantial deficits in vehicle control relevant in real-world situations. Operational outcomes showed particularly strong effects of acute sleep deprivation, with a surprisingly rapid deterioration of lateral control. SDLP increased by 7.86 cm on average—a 53% rise from Module 1 to Module 3 in condition D—emerging within 4–6 minutes and continuing over the full 12-minute drive. The magnitude of this deterioration is comparable to the impairment reported at a blood alcohol concentration of approximately 1.2–1.4 g/L (differential increase in SDLP between C and D in Module 3: 5.96 cm, +35.19%) (27), and aligns with previously observed increases immediately prior to falling asleep at the (simulated) wheel (1).

Such early, progressive, and substantial impairment, occurring in the absence of corresponding declines in vigilance performance, suggests that drivers may substantially underestimate, or even fail to perceive, how severely sleep loss compromises their operational control.

This also suggests that a driver’s decision to undertake even a short highway drive after 24 hours of wakefulness may be based on an overestimation of functional capability. Under these conditions, a driver may not be able to reach the next highway exit before experiencing dangerously reduced vehicle control or even a sleep-related accident (1) triggered by a microsleep event.

Despite the strong implications and real-world relevance, the present study has several limitations. First, naturalistic studies using driving simulation allow only an indirect approximation of real-world accident risk and therefore cannot be directly translated to real-life driving outcomes. Second, the time-on-task analysis was restricted to a short, 12-minute monotonous drive, and the tone-based vigilance task may have been too brief or insufficiently sensitive to detect subtle effects of the sleep-loss regimes. Third, the primary aim of the overarching study - metabolomic pattern recognition of sleepiness - necessitated an initial focus on a cohort of young, healthy men, which limits generalizability to the broader population. At the same time, this cohort represents a demographic subgroup with one of the highest risks for drowsy driving and sleep-deficit–related accidents (28), underscoring the relevance of these findings for targeted countermeasures, regulatory consideration of sleep-related impairment in road safety policies and public awareness.

Independent of which specific preventative measures may ultimately prove most effective in reducing sleepiness-related traffic fatalities, a combined approach is likely required.

This includes technological interventions, such as advanced driver assist systems that monitor and warn for signs of sleepiness, improved tools to assess the degree of sleepiness or impairment, and the communication of clear, tangible evidence from human-centered research on the effects of sleep loss related to alcohol as demonstrated here and elsewhere (1, 27), highlighting the serious risk of drowsy driving on the road.

Beyond such measures, the present findings also highlight challenges in translating scientific evidence on sleep-related impairment into regulatory frameworks. In this context, it is noteworthy that some jurisdictions have begun to formally recognize prolonged wakefulness as a risk factor for impaired driving. For example, “Maggie’s Law” in the U.S. state of New Jersey (29) classifies driving after more than 24 hours of continuous wakefulness as reckless driving. While such legal approaches remain rare and controversial, they underscore growing recognition that extended wakefulness can impair driving performance to a degree comparable to elevated blood alcohol concentrations, as shown in the present and previous studies (1, 23), and illustrate the need for clear, evidence-based communication of sleep-loss–related driving risk.

## Supporting information

Supplemental Methods and Results

## Data Availability

All data produced in the present work are contained in the manuscript

## Acknowledgements

The authors would like to express profound gratitude to all persons involved in this study.

## Author Contributions

SL established the driving simulator, contributed to the conceptualization of the driving experiment, supervised all driving simulation experiments and performed analyses in R, re-confirmed statistical analyses in GraphPad Prism, established figures and wrote/revised the manuscript. MS was responsible for trial management, contributed to the conceptualization of the driving experiment, and led the data collection of the overarching study. TK was the sponsor and co-initiated the project. HPL was study site leader and co-initiated the project. KK was principal investigator. All authors contributed to the design and development of the overarching trial, discussed the data, read and approved the final manuscript.

## Data Availability

Data, rationale, code, and detailed results of all mentioned and additional analyses are provided as .html files as R-Markdown in the online supplemental material.

