## Supplemental Methods and Results for "Effects of acute or cumulative 8-hour sleep loss on simulated driving in a naturalistic, monotonous night-time setting: a randomized, controlled, crossover trial"

[1] Department of Traffic Medicine, Zurich Institute of Forensic Medicine, University of Zurich, Zurich, Switzerland, [2] Department of Forensic Pharmacology and Toxicology, Zurich Institute of Forensic Medicine, University of Zurich, Zurich, Switzerland, [3] Institute of Pharmacology & Toxicology, University of Zurich, Zurich, Switzerland, [4] Sleep & Health Zurich, University of Zurich, Zurich, Switzerland

[*] to whom correspondence should be addressed: Stefan Lakämper, Institute for Forensic Medicine, Division of Traffic Medicine, University of Zurich, Zurich, Switzerland, Andreasstrasse 15, 8050 Zurich, Switzerland,

### Detailed Methods

#### Driving Simulator Setup

The driving simulator VICTOR (Vehicle for Interdisciplinary Clinical and Translational Research) has been described previously (1). The simulator is based on a full BMW i3 chassis, in which each wheel's shock absorber was replaced by spindle actuators (Festo, Esslingen, Germany; actuator ESBF-BS-50-100-20P, servomotor EMMB-AS-80-07-S30S, control servo drive CMMT-AS-C4-3A-PN-S1; Neugart, Kippenheim, Germany, gear Art-No. 58474) using partially custom-built components. This configuration provides motion cues primarily reflecting road surface imperfections, acceleration, and braking.

Side-view mirrors and the dashboard monitor were replaced with LCD displays (Beetronics 10HD7 or 13HD7). Rear-view information was presented on a 40-inch monitor (IIYAMA X4071UHSU-B1) mounted in the trunk. The central console featured a touchpad (Asus Zen Touch MB16AMT) for displaying and collecting additional task-related information. Controls for throttle, brake, indicator, and steering were modified to enable integration into the virtual driving environment.

Vehicle dynamics simulation and virtual environment generation were provided by SILAB (version 7.0; WIVW, Würzburg, Germany). Visual content was displayed either within the vehicle (mirrors) or projected via five DLP laser-phosphor projectors (Barco F-80_Q7, optics GLD 0.85–1.06:1), mounted on custom position controls (Lumachroma, Switzerland), onto a 275° stable drywall projection barrel. Five images at a resolution of 2560 × 1600 pixels (WQXGA) were mapped, warped, and blended using VIOSO-AnyBlend

(Vioso, Düsseldorf, Germany). Both visual display and recording of driving parameters were performed at a sampling rate of 60 Hz.

The overall system was controlled by SILAB via a single LAN-operated workstation coordinating ten identical computers (Intel Core i7-9700 @ 3.6 GHz [8 cores], 16 GB RAM, NVIDIA GeForce RTX 2080 SUPER). The facility was air-conditioned to maintain an ambient temperature of approximately 22 °C (range: 20–24 °C). The vehicle interior was supplied with fresh air at 21 °C via the building's ventilation system. Technical equipment and control hardware were concealed within the trunk or otherwise kept out of sight, resulting in a minimalistic appearance closely resembling a vehicle parked in a garage and providing a low-threshold, realistic experimental setting for participants. A virtual 360° tour of the simulator setup is available online (2).

#### Simulated Driving Task and Data Acquisition

The simulated driving task consisted of a set of standardized scenarios that have been previously validated and specifically adapted for Swiss roads for the assessment of driver fitness (SPCH-DFA; WIVW, Würzburg, Germany). Several scenarios were repeated within the task, in some cases with minimal modifications. For clarity, each scenario is hereafter referred to as a *module* and is described according to its sequence within the experimental procedure.

The simulated driving assessment comprised a total of twelve modules. Modules 1–3 each consisted of an approximately four-minute, highly monotonous, night-time car-following task. Participants were instructed to follow a lead vehicle at an appropriate distance on a single-lane highway, with overtaking strictly prohibited. The lead vehicle traveled at a speed fluctuating between 80 and 90 km/h (speed limit: 100 km/h), while oncoming traffic was simulated at irregular intervals of 5–20 seconds (see Video S1 in (1)).

To minimize external stimulation, induce soporific conditions, and promote sustained focus on the tracking task, the simulator's speedometer was deactivated during designated rating phases, thereby reducing light emission within the cockpit. Road surface irregularities and changes in vehicle speed or position were conveyed via the motion system. However, no additional motion cues were provided in the event of collisions or lane departures. Instead, leaving the roadway resulted in the vehicle contacting an invisible "digital wall" located approximately 10 meters from the road edge, without any visual or auditory feedback to the participant (Video S2 in (1)).

Modules 1–3 additionally included a tone-based vigilance task. Drivers heard a regular sequence of tones presented at 3,400-ms intervals and were instructed to press a button located on the left indicator stalk whenever a tone was omitted (3).

Module 4 served as a transition phase, during which the lead vehicle departed, the dashboard displays were reactivated, and a tunnel drive was announced. Module 5 consisted of a brightly illuminated, winding tunnel, after which environmental conditions transitioned from night-time to a slightly foggy, early-morning rural setting. Modules 6–9 included additional traffic-related events, such as wooded road segments, an overtaking vehicle (modules 6–8), and a police car approaching from behind (module 9). In Module 10, the police vehicle signaled the participant to pull over at a designated bus stop, as previously instructed, and participants were informed that a routine traffic control would follow. Provisionary Modules 11 and 12 included escalated auditory and visual cues prompting the driver to stop in case the initial signal in Module 10 was missed, culminating in a forced stop scenario.

Both vigilance-related and driving performance parameters were recorded using the SILAB system and analyzed post hoc. Standard driving errors, including lane departures, near collisions, and collisions, were additionally tracked using the SAFE software. The total duration of the evaluated driving task (Modules 1–9) was approximately 20 minutes (mean duration: 19:26 minutes).

**Study Procedure and Design**

The full study protocol was pre-registered and published (3). For the primary outcome—changes in the oral fluid metabolic profile (reported elsewhere)—a minimum sample size of 17 participants was determined a priori. A simulated drive conducted during a screening visit, encompassing Modules 1–7, is reported here in part as baseline data (condition B) in Supplemental Figure 1.

Prior to the experimental study drives, twenty healthy young men, each habitually sleeping approximately eight hours per night, completed three experimental conditions: normal sleep (condition C) or one of two sleep-loss interventions: (1) acute sleep deprivation (condition SD), achieved by skipping one entire night of sleep (8-h deficit), and (2) cumulative sleep restriction (condition SR), achieved by curtailing sleep by two hours per night over four consecutive nights (4 × 2-h deficit). Both interventions resulted in an equivalent total sleep loss of eight hours.

The study employed a randomized, cross-over design. Adherence to the protocol and behavioral instructions was closely monitored by the study team in a controlled sleep-laboratory environment. Time in bed during home sleep periods was verified using actigraphy (GENEActiv, Activinsights Ltd, Kimbolton, UK) and daily sleep–wake diaries. Before each simulated drive, subjective sleepiness was assessed using the Karolinska Sleepiness Scale (KSS, version A).

**Data analysis**

Driving and vigilance data were analyzed for modules 1-9 according to comparable studies on sleepiness and driving (1, 4). All analysis steps were implemented using R (R Foundation for Statistical Computing) and documented using R Markdown to ensure full reproducibility. Rendered analysis reports and resulting tables are provided as HTML files in the online Supplemental Material (NS01–NS03).

In brief, raw data were aggregated and harmonized, and measurement units were introduced or converted as required to generate a standardized base dataset. Driving and vigilance parameters were subsequently computed at both the drive level and the module level. These included the standard deviation of lateral position (SDLP), speed (SDS), and distance to the lead vehicle (SDD), as well as measures of inappropriate line crossings (ILC), including total count (tcILC), total duration (ttILC), and average duration (atILC). Analogous metrics were derived for parallel danger (PD) events and collisions. The two-level PD parameter quantified the lateral proximity of the ego vehicle to oncoming traffic, distinguishing between standard and reduced lateral distances as indicators of increasing collision risk. Because PD events and collisions were extremely rare, these parameters were not included in the present analyses. Specifically, only one collision occurred during experimental conditions and one during screening (P026–SD–M3 and P004–B–M6, respectively).

Steering control was quantified using counts of steering wheel reversals (cSWR), defined as events in which steering velocity reversed direction by at least ±3°/s within a 1-s interval. Vigilance performance was assessed using counts of Tone, NoTone, and Signal events, as well as hits, misses, false positives, and correct rejections. Sensitivity, specificity, and mean reaction time to hits were calculated using standard definitions.

For visualization and data inspection, summarized datasets were exported to GraphPad Prism. Screening data were excluded from inferential analyses. Outliers were identified, evaluated, and excluded only when clearly justified. Rather than applying data transformations, isolated outlying observations were removed on a per-parameter basis, as only a very small number of values met exclusion criteria. In total, five drive-level observations (SDLP–P026–SD; SDS–P010–SD; SDS–P021–C; SDD–P017–SD; tcILC–P026–SD) and one module-level observation (SDD–P017–SD–M1) were excluded.

For both drive-level and module-level analyses, parameter values were standardized using z-scores. Integrated driving scores (IDS) were then computed by summing standardized parameters and re-standardizing the resulting composite score, using drive-level variables SDLP, SDS, SDD, tcILC, ttILC, atILC, sensitivity, reaction time, and cSWR. For module-level IDS (IDS_M), a reduced parameter set (SDLP, SDS, tcILC, ttILC, atILC, cSWR) was used to ensure comparability across modules. Parameters unavailable for specific modules were treated as zero (i.e., average performance), preventing distortion of composite scores. All summarized IDS data are provided in the Supplemental Material.

Drive-level outcomes were analyzed descriptively and inferentially using a stepwise modeling approach that progressively accounted for the hierarchical data structure. Initial fixed-effects ANOVA models including Condition, Participant, and Visit were used for orientation only. Linear mixed-effects models were then applied, with Participant modeled as a random intercept, followed by a final model additionally including Visit as a random intercept. This specification was considered the primary model, as it appropriately accounts for repeated measures within participants and visits and supports generalization beyond the observed sample.

Linear mixed-effects analyses (Model 3) were conducted for Modules 1–3 to assess time-on-task effects during identical driving segments. These analyses identified condition-by-module interactions for operational driving parameters, including SDLP and the module-level integrated driving score (IDS_M). Detailed model outputs and pairwise contrasts are provided in the Supplemental Files (20250729_module_level_LMMfull.xlsx)

### Supplemental Results

#### Screening baseline comparison

A comparable simulated driving task has previously been assumed - and qualitatively shown - to be accessible without extensive familiarization (1).

The simulated driving task in this study was practiced once during the screening visit (condition B, module 1-7). The simulated driving task in this study was practiced once during the screening visit (condition B; Modules 1–7). None of the screened participants reported pronounced symptoms of simulator sickness.

Using a simple mixed-effects analysis ($F(3, 50) = 9.578$, $p < 0.0001$; total df = 73; GraphPad Prism), applied to the lateral control parameter SDLP across all available drives (B = 24, C = 19, SR = 18, SD = 18), we quantitatively confirmed that no significant difference was present between mean SDLP values in conditions B and C (B vs. C mean difference = 0.37 cm [−2.78 to 3.51], adjusted p = 0.9897). This finding indicates the absence of a measurable learning or familiarization effect for this task.

We therefore conclude, on a quantitative basis, that this and comparably simple simulated driving tasks (i.e., without complex urban scenarios) can be employed without prior familiarization, at least in cohorts of participants who report no difficulties with simulator exposure, virtual reality headsets, amusement park rides, or sitting backwards on a train.

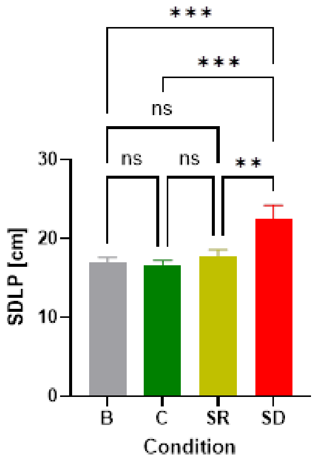


**Figure S1.** *Mean standard deviation of lateral position (SDLP) across conditions.* Bars show mean SDLP (± SEM) for the screening drive (B), normal sleep (C), cumulative sleep restriction (SR), and acute sleep deprivation (SD). No significant difference was observed between conditions B and C, indicating no measurable familiarization effect. Acute sleep deprivation (SD) resulted in significantly increased SDLP compared with all other conditions ($p < 0.01$–$0.001$); ns = not significant.

Furthermore, the overall mean SDLP values obtained in conditions B, C, and SR (16.85, 16.47, and 17.77 cm, respectively) are comparable to—if not identical with—SDLP values reported at 0 g/L blood alcohol concentration (BAC) in on-road naturalistic driving studies (5). These reference values, together with SDLP values at higher BACs, have previously been used (1) to contextualize the effects of sleepiness relative to alcohol, albeit based on relative differences in a mixed cohort of potentially sleepy participants. The close correspondence of SDLP values obtained here from healthy male participants under strictly controlled conditions (rested state, sleep laboratory, polysomnography, 0 g/L BAC) with those reported in (5) provides a robust naturalistic reference, allowing the effects of acute sleep deprivation (condition SD) to be interpreted on a sound and ecologically grounded basis (see Discussion).
